# Cilostazol may be a better therapeutic agent for acute ischemic stroke because of its efficacy in preventing post-stroke depression

**DOI:** 10.1101/2023.10.30.23297771

**Authors:** Yuling Zhang, Ganggang Yang, Rui Wang, Xin Wang, Xiying Tan, Chaozhi Tang

## Abstract

Post-stroke depression (PSD) has more than 1/3 complications in ischemic stroke (IS) patients, but there is still no specific drug that can effectively prevent the occurrence of PSD. Cilostazol, commonly used in the treatment of IS, has been found to rescue cognition impairment in recent years, but whether it has the efficacy in preventing PSD remains uncertain. In this study, a total of 431 acute ischemic stroke (AIS) patients treated with and without cilostazol during 2021-2022 were recruited and followed up for 6 months. At the end of follow-up, we found that AIS patients of cilostazol-treated group had significantly lower National Institutes of Health Stroke Scale (NIHSS) and 24-item Hamilton’s Depression Scale (HAMDS) scores than patients of cilostazol-free group (*p*<0.05), the total incidence of depression was 15.47% in patients of cilostazol-treated group and 36.02% in patients of cilostazol-free group, including 8.84% and 16.67% mild depression (*p*<0.05), 6.08% and 12.90% moderate depression (*p*<0.05), 0.55% and 6.45% major depression (*p*<0.01), respectively. In addition, the plasma levels of high-sensitive C-reactive protein (Hs-CRP), homocysteine (HCY), tumor necrosis factor-α (TNF-α), interleukin-1β (IL-1β) and neutrophil/lymphocyte ratio (NLR) in AIS patients of cilostazol-treated group were all lower than those in patients of cilostazol-free group (*p*<0.05). These results suggest that cilostazol is a better therapeutic agent for AIS because it may prevent PSD by decreasing the levels of inflammation in patients.

## Introduction

Ischemic stroke (IS) is an acute disease of cerebrovascular circulation disorder, and which can lead to death when vascular infarction is severe. More than one third of IS patients were complicated with post-stroke depression (PSD), showing more serious clinical characteristics such as delayed consciousness, memory loss, language and cognitive disorders, sleep disorders, emotional depression, difficult treatment and strong suicide tendency than IS patients alone, which greatly reduces the recovery rate of patients[1, 2]. As drugs that regulate neural activities such as consciousness, cognition or emotion inevitably have some side effects on the human brain, IS patients may have more severe adverse drug event when taking them, it is not recommended to apply additional antidepressant drugs to IS patients to prevent the occurrence of PSD. For example, the results of two large clinical studies have shown that early administration of fluoxetine to stroke patients can reduce the incidence of PSD, but does not improve the functional outcome. Instead, it increases the risk of falls, bone fractures, epileptic seizures and hyponatremia[3, 4]. There is no international report on effective and rational drugs or methods for PSD prevention and can be implemented internationally. Therefore, the prevention of PSD has become a bottleneck limiting the cure rate of IS patients.

Cilostazol, a specific phosphodiesterase (PDE)-3 inhibitor, which can block cAMP degradation, inhibit adenosine uptake, platelet aggregation and dilate blood vessels, has been used in the treatment of atherosclerosis, intermittent claudication, IS and other vascular embolic diseases[5–7]. With the continuous in-depth study on the efficacy of cilostazol, it has also been found to regulate the level of inflammation in the body, for example, by inhibiting the activation of nuclear factor-κB (NF-κB) to protect the heart injury caused by hypercholesterolemia[8]. Studies on mouse models of atherosclerosis have also shown that cilostazol can significantly improve the proinflammatory state of vascular endothelial cells and slow down the progression of atherosclerosis by down-regulating intercellular adhesion molecule-1 (ICAM-1), monocyte/macrophage-2 (MOMA-2), tumor necrosis factor-α (TNF-α), interleukin-6 (IL-6) and other inflammation-related factors[9]. In limb ischemic model mice, cilostazol increases the levels of IL-4, IL-10, IL-6 and IFN-γ, and decreases the expression of IL-2 and inflammatory cell infiltration[10].

In recent years, studies have shown that PSD or depression is closely related to the body inflammation levels change[1, 11–16]. There is an acute inflammatory response after a stroke, involving increased levels of pro-inflammatory, both centrally and peripherally. In some cases, the adaptive acute response does not resolve and leads to chronic inflammation, a detrimental phenomenon[1]. Abnormal inflammatory response or inflammatory index, such as high-sensitive C-reactive protein (Hs-CRP), interleukin-1β (IL-1β), TNF-α, IL-6, systemic immune-inflammation index (SII), neutrophil-to-lymphocyte ratio (NLR), platelet-to-lymphocyte ratio (PLR) and derived NLR (dNLR), have been linked to PSD or depression[13–16]. Villa et al. hold that, in the future, especially in patients affected by inflammation processes, novel developments might point at anti-cytokine modulators which can improve symptoms of depression[17].

Cilostazol was also try to antidepressant treatment. A double-blind, randomized, placebo-controlled clinical study of 80 depressed patients with HAMDS scores of 20 or more from Egypt showed that cilostazol significantly alleviated depression after 6 weeks of treatment and was superior to placebo in the improvement of detected neurotrophic and inflammatory biomarkers[18]. Similar findings were also confirmed in a parallel randomized controlled trial of depressed patients in Iran[19]. These clinical analyses indicate that cilostazol is safe and effective for the treatment of depression. In addition, clinical treatment cases from Japan reported that 7 AD patients with depression, 1 PSD patient with depression, and 2 elderly patients with major depressive disorder were significantly relieved after taking cilostazol[20–22]. Kim et al. applied cilostazol to intervene PSD model mice, and found that cilostazol could significantly alleviate the depressive behavior, inhibit microglial activation and reduce neuronal death in hippocampus region[23].

The combined effect of cilostazol on vascular disease, inflammation and depression suggests that cilostazol may play a preventive role in the development of PSD through inflammatory regulatory pathway. According to the above clinical research, case reports and animal experiments, cilostazol does have a good preventive effect on PSD patients. However, the clinical sample size supporting this view is too small. On the basis of these findings and our clinical experience, it is necessary to conduct scientific exploration and fully demonstrate the efficacy and mechanism of cilostazol in preventing PSD. Complete analysis cilostazol potential preventive effect on PSD will provide a new perspective for clinical prevention and treatment of PSD. To this end, 431 acute ischemic stroke (AIS) patients who were hospitalized were recruited and followed up for 6 months to evaluate the preventive efficacy of cilostazol on PSD and its possible mechanisms.

## Methods

### Study population

The subjects were first episode AIS patients who were hospitalized at Xinxiang First People’s Hospital during the period from 1 January 2021 to 31 December 2022. Patients admitted to the hospital within the first 24 h after stoke onset were consecutively recruited and followed up for 6 months. AIS was defined according to the World Health Organization Multinational Monitoring of Trends and Determinants in Cardiovascular Disease criteria and verified from computed tomography (CT) or magnetic resonance imaging (MRI) reports performed within 24 h after admission in all patients. The subjects should also have the ability of word recognition, be able to understand the survey content, have no cilostazol allergy and antiplatelet therapy contraindications, and have no surgical plan during the follow-up period, have no received psychiatric treatment or taken antidepressant or anticoagulant drugs within 4 weeks before admission. Exclusion criteria were a patient self-report of having been hospitalized for any psychiatric illness, pre-stroke diagnosis of dementia or cognitive impairment, decreased level of consciousness, severe aphasia or dysarthria, metabolic abnormalities, liver insufficiency, acute medical illness (e.g., infection, autoimmune disease, malignant tumor), and acute neurological illness other than stroke. A total of 431 patients aged 37-84 years were enrolled in this study, of which 215 received conventional AIS treatment (cilostazol-free group) and 216 received conventional AIS treatment supplemented with 200 mg/d cilostazol (H10960014, Zhejiang Dazhong Pharmaceutical Co., LTD.) (cilostazol-treated group).

### Examinations at admission

Baseline data were collected at admission, including age, gender, body mass index (BMI), smoking (continuous or cumulative smoking history≥6 months, smoking≥1 cigarette/day), history of heavy drinking (≥5 standard drinks/day), hypertension, hyperlipidemia (plasma total cholesterol ≥6.2 mmol/L and/or plasma triglycerides≥2.3 mmol/L), history of diabetes and heart disease, lesion location (cortex, subcortical, cerebellum and brainstem), etc. The National Institutes of Health Stroke Scale (NIHSS) and 24-item Hamilton’s Depression Scale (HAMDS) scores and plasma levels of (Hs-CRP), homocysteine (HCY), TNF-α, IL-1βand NLR were also measured at admission.

### Follow-up and post-treatment examinations

Since PSD symptoms such as cognitive impairment and emotional disorder in AIS patients generally appeared within 6 months of stroke onset[1, 24, 25], each enrolled patient was followed up for 6 months in this study. According to the recovery degree of AIS, the hospital discharge time and medication time were evaluated by the doctor in charge, and the follow-up time points of PSD remained unchanged. For patients who actively withdrawal or developed depression within 6 months and were treated with other anti-depression drugs, the diagnosis time of depression was taken as the end point of follow-up for post-treatment examination. At the end point of follow-up, NIHSS and HAMDS scores and plasma levels of Hs-CRP, HCY, TNF-α, IL-1β and NLR were examined. Face-to-face adherence reminder will take place at every treatment session. Adherence to the protocol as provided in the informed consent form is assessed at every follow-up session. This protocol is conducted according to the 2013 Standard Protocol Items: Recommendations for Interventional Trials (SPIRIT) guidelines[26]. The assessment schedule, including eligibility assessments, is outlined in Fig 1.

**Figure 1.**
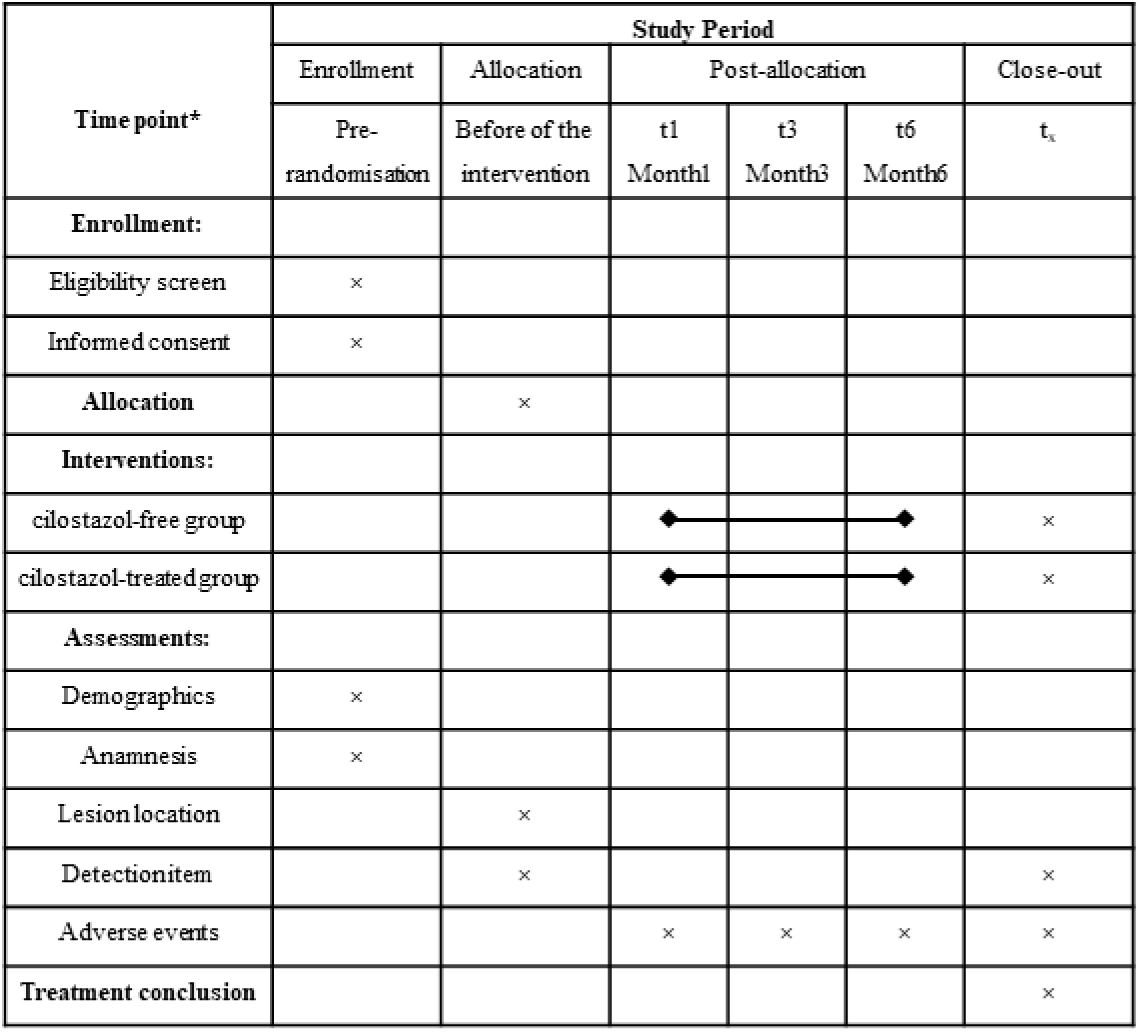
SPIRIT schedule of enrolment, interventions and assessments. t**_x_:** The diagnosis time of depression was taken as the end point of follow-up.

### NIHSS and HAMDS scores

The NIHSS score ranged from 0 to 42, and the higher the score, the more severe the nerve injury. 24-item HAMDS score criteria were as follows: score<8 was judged as no depressive symptoms, 8≤score<20 was judged as mild depressive symptoms, 20≤score<35 was judged as moderate depressive symptoms, score≥35 was judged as major depressive symptoms. The NIHSS cores were assessed by two trained and consistent assessors, who were unaware of the patient’s group and medication status. Both scale assessors participated in standardized training before the program start. The assessors and the training physician conduct NIHSS assessments on stroke patients within a specified timeframe. The assessor’s NIHSS scores for each item are statistically analyzed and compared with the training physician’s scores to evaluate consistency. If the results show no significant difference, the assessor is considered to have passed the training. The NIHSS scoring results take the mean of the two assessors. The NIHSS and HAMDS score results were again reviewed by another assessor to avoid calculation errors.

### Blood collection and laboratory test

Fasting blood samples were collected intravenously via venipuncture in Vacutainer^®^ PPT^TM^ Plasma Preparation Tube (362800, Becton Dickinson, San Jose, CA, USA) at 8:00 am. Some samples were classified and counted by blood cell analyzer (UniCel DxH800, Beckman Coulter), and the NLR value was calculated. The other samples were centrifuged at 1000× g for 10 min, and the plasma was separated and stored at -80 ℃, the concentration of biomarker molecules were respectively determined by Hs-CRP (AB260058, Abcam), HCY (HLE10135, Shanghai Haring Biotechnology Co., LTD), TNF-α (AB181421, Abcam) and IL-1β (AB214025, Abcam) enzyme-linked immunosorbent assay kits. For all measurements, levels that were not detectable were considered to have a value equal to the lower limit of detection of the assay.

### Sample size

Sample size calculations have been described previously[27]. The HMADS score of the subjects was the main outcome index. According to a previously performed pilot study and published paper[28]. The standard deviation was 5.66, a two-tailed significance level of 5%, a statistical power of (1-β) was 0.8, and the sample size ratio between the cilostazol free group and cilostazol treated group was 1:1. Making allowance for a drop-out rate of 20%, 68 patients were needed in each group.

### Ethics

Written informed consent was obtained from all patients or authorized representatives prior to testing, and this study conformed to the principles of the Declaration of Helsinki and was approved by the Ethics Review Committee of Xinxiang First People’s Hospital (approval No:2021010402), and has been registered at https://www.chictr.org.cn/. In the informed consent form, we explained the nature of the trial, the purpose, the possible benefits and risks, other alternative treatments, so that the subjects can fully understand and express their intentions to eliminate potential conflicts of interest. A model consent form is also available from the corresponding author on reasonable request.

### Randomization and blinding

A random permuted block randomization list with a 1:1 allocation ratio was generated on a computer using statistical software (SPSS, IBM version 26) by an independent researcher who were not involved in treatment, scale evaluation, and experimental testing. To hide the grouping information, the grouping form was placed in an opaque sealed envelope with the patient number marked outside. The envelopes were kept by the study coordinator, who opened them and informed the clinician before using cilostazol. Patients, scale assessors, and experimental testing personnel were blinded to treatment allocation. Potential confounding factors include age, sex, body weight, blood pressure, blood lipids, infarction location and so on. We used the above randomized method and blind method to eliminate the impact of potential confounding factors on the outcome events.

### Data collection and management

Our clinical research team develops a data security monitoring plan based on risk. All clinical observations will be recorded in CRF (case report form, CRF) in a timely manner. The patient’s original CRF, informed consent form, clinical history, original test sheet and raw data must be traceable. The designated researcher must manually complete the CRF, regularly collect the necessary quality control and safety evaluation information, and use the double-entry method to digitize the data. The research assistant will check the original subject records, mark the data in question, and ask the relevant personnel to confirm or correct them. Access to this data will be limited to specific research teams.

### Statistical analysis

We assigned patients equally into cilostazol-free group and cilostazol-treated group by block randomization method to reduce errors resulting from confounding factors such as temperature change, age, gender, educational background, and patients’ own underlying diseases such as hypertension and hyperlipidemia, and so on. Significance analysis of baseline data of general clinical data and test data at the end of follow-up by the t test, Mann-whitney *U*-test or Fisher’s exact test. Kolmogorov-Smirnov normality test was used to detect whether the measurement data of each group conformed to the normal distribution, and the measurement data in accordance with the normal distribution were expressed by mean±standard deviation (M±SD). The differences between the two groups were compared by parameter t-test. The metrological data that do not conform to the normal distribution are represented by the median and quartile spacing M (Q1 and Q3). Mann-whitney *U* test was used to analyze the significance. The counting data were expressed by frequency and percentage, and the significance was compared by Fisher exact test. Spearman correlation analysis was used to analyze the correlation and causal relationship between depression incidence and NIHSS score. Because there is no significant difference in the baseline level of general clinical data (confounding factor) between the two groups, the correlation analysis of HAMDS score and NIHSS score is only a simple linear regression, and there is no need to adjust the interference of other confounding factors. R represents correlation coefficient, *p* value represents whether the correlation coefficient R of NIHSS score and HAMDS score is statistically significant. *p* value<0.05 indicates that the correlation coefficient R has statistically significant significance; *p* value<0.01 indicates that the correlation coefficient R has extremely significant statistical significance. Positive correlation between them is R>0, and vice versa. Outcome analyses were performed in the modified intention-to-treat population, which included all randomly assigned patients except those who withdrew consent for participation in the trial and those lost to follow-up. No imputation of missing data was conducted. All the analyses and drawings were carried out using GraphPad Prism 8.0 software. In the t test, Mann-whitney *U*-test or Fisher’s exact test and so on, two tailed 95% confidence intervals *p* value<0.05 were considered statistically significant, *p* value<0.01 were considered extremely statistically significant (\**p*<0.05, \*\**p*<0.01).

## Results

### Basic characteristics of patients at admission

In the cilostazol-free group, 29 patients withdrew from the study due to adverse drug event (n=9), hospital transfer (n=8) or death (n=14, including 2 patients with adverse drug event), and 186 patients completed follow-up and analysis. In the cilostazol-treated group, 35 patients withdrew from the study due to adverse drug event (n=12), hospital transfer (n=19, including 2 patients with adverse drug event) or death (n=7, including 1 patient with adverse drug event), and 181 patients completed follow-up and analysis. We compared basic characteristics at admission between two groups, there were no significant differences in age composition, sex ratio, adverse living habits (such as smoking and alcoholism), past medical history, lesion location, NIHSS and 24-items HAMDS scores, plasma levels of Hs-CRP, HCY, TNF-α, IL-1β and NLR (*p*>0.05) (Tab. 1). These basic data indicate that follow-up findings were comparable between the two groups of patients. In addition, according to the HAMDS scores of the two groups at admission, no depressive symptoms occurred in the subjects, which is suitable for the demonstration of the preventive efficacy of cilostazol on PSD.

**Table 1.** Baseline characteristics of patients.

|  | Cilostazol-free group<br>(n=186) | Cilostazol-treated group<br>(n=181) | *p* value |
| --- | --- | --- | --- |
| Age (IQR, years) | 69.00 (63.00, 75.25) | 70.00 (63.00, 74.00) | 0.696 |
| Female, n (%) | 81 (43.55) | 73 (40.33) | 0.597 |
| BMI (kg/m<sup>2</sup>) | 25.35 ± 3.04 | 24.87 ± 2.79 | 0.099 |
| Active smokers, n (%) | 34 (18.28) | 37 (20.44) | 0.692 |
| Heavy alcohol consumption, n (%) | 9 (4.84) | 5 (2.76) | 0.415 |
| Anamnesis |  |  |  |
| Hypertension, n (%) | 107 (57.53) | 95(52.49) | 0.346 |
| Hyperlipidemia, n (%) | 66 (35.48) | 58 (32.04) | 0.509 |
| Diabetes, n (%) | 41 (22.04) | 46 (25.41) | 0.464 |
| Heart disease, n (%) | 24 (12.90) | 28 (15.47) | 0.550 |
| Mental disorders, n (%) | 5 (2.69) | 6 (3.31) | 0.768 |
| Lesion location |  |  |  |
| Cortex, n (%) | 109 (58.60) | 118 (65.19) | 0.199 |
| Basal ganglia, n (%) | 75 (40.32) | 83 (45.86) | 0.294 |
| Thalamus, n (%) | 63 (33.87) | 71 (39.23) | 0.329 |
| Cerebellum, n (%) | 10 (5.38) | 6 (3.31) | 0.445 |
| Brainstem, n (%) | 14 (7.53) | 7 (3.87) | 0.177 |
| Detection item |  |  |  |
| HAMDS score |  |  |  |
| Asymptomatic (<8), n (%) | 186 (100) | 181 (100) | >0.99 |
| Mild (8~19), n (%) | 0 (0) | 0 (0) | >0.99 |
| Moderate (20~34), n (%) | 0 (0) | 0 (0) | >0.99 |
| Major (≥35), n (%) | 0 (0) | 0 (0) | >0.99 |
| NIHSS score (IQR) | 12 (7, 16) | 12 (8, 16) | 0.959 |
| Hs-CRP (IQR, mg/L) | 12.97 (10.98, 15.03) | 12.64 (9.92, 15.63) | 0.219 |
| HCY (IQR, μmol/L) | 18.38 (15.85, 21.69) | 18.62 (16.92, 20.64) | 0.315 |
| TNF-α (IQR, ng/L) | 37.68 (34.01, 41.64) | 37.43 (33.98, 40.76) | 0.341 |
| IL-1β (IQR, ng/L) | 46.70 (40.73, 50.52) | 44.82 (41.49, 49.30) | 0.199 |
| NLR (IQR) | 3.35 (2.44, 4.07) | 3.35 (2.76, 4.27) | 0.375 |

Note: The measurement data in accordance with the normal distribution were expressed as the mean±standard deviation, and those that do not conform to the normal distribution were expressed as the median (the distance between the upper and lower quartiles). Enumeration data were given in the form of a count (percentage).
Abbreviations: *IQR* interquartile range, *BMI* body mass index, *n* the number of people.

### Incidence of adverse drug event during follow-up

We statistically analyzed the incidence and differences of adverse drug event between the two groups during the follow-up period. The results showed that the incidence of adverse drug event in cilostazol-treated group was 4.19%, and that in cilostazol-free group was 5.56%, and there was no significant difference between them (*p*=0.656), indicating that cilostazol treatment did not increase the incidence of adverse drug event.

### NIHSS and HAMDS scores

NIHSS and HAMDS scores were assessed at admission and at the end of follow-up. The results of NIHSS scores showed that there was no significant difference in the severity of AIS between the two groups at admission (*p*=0.959), however, at the end of follow-up, the NIHSS scores of cilostazol-treated group was significantly different from that of cilostazol-free group (*p*=0.036) (Tab. 1 and Fig. 2). Moreover, the dispersion of NIHSS scores in patients of cilostazol-treated group was smaller than that in cilostazol-free group (Fig. 2). These results suggest that the clinical use of cilostazol is very beneficial to the recovery of neurological deficits in AIS who have no contraindication history to cilostazol. Cilostazol treatment has a better therapeutic efficacy on AIS and can increase the prognosis stability of AIS.

**Figure 2.**
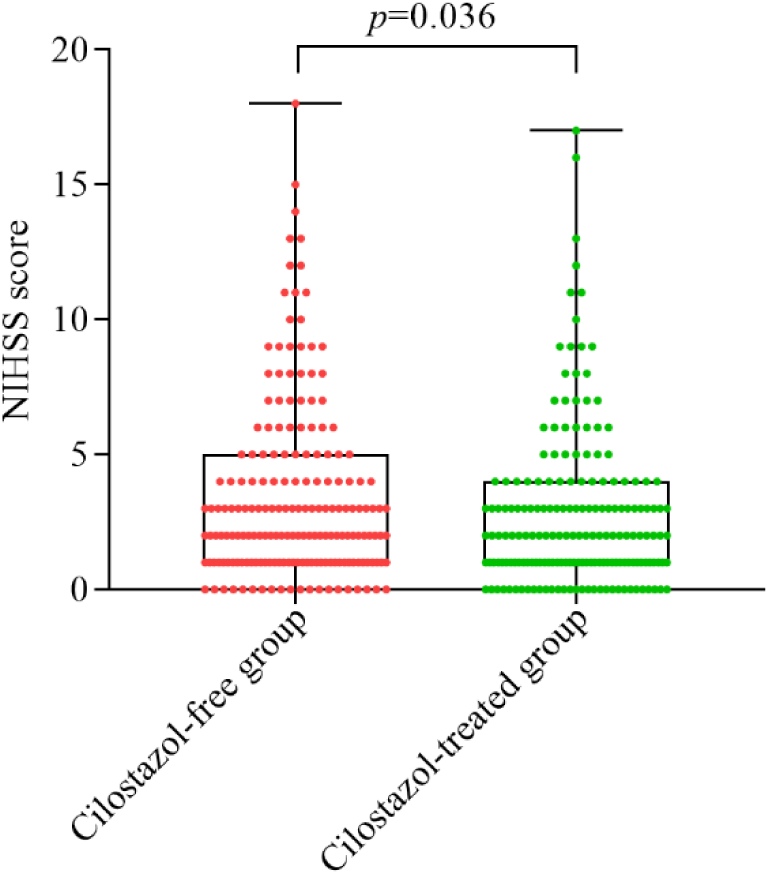
NIHSS scores in patients of the cilostazol-free group and cilostazol-treated group at the end of follow-up. Mann-Whitney *U* test was used to analyze whether there was significant difference in NIHSS score between the two groups.

HAMDS scores showed that there were no depressive symptoms in both groups at admission, but PSD was found in both groups at the end of follow-up, and the incidence of PSD in cilostazol-treated group (15.47%) was significantly lower than that in cilostazol-free group (36.02%) (*p*<0.0001). From the perspective of depressive symptoms grade, the incidence of mild PSD (8.84% vs 16.67%, *p*=0.032) and moderate PSD (6.08% vs 12.90%, *p*=0.029) in cilostazol-treated group were significantly lower than those in cilostazol-free group, and the incidence of major PSD (0.55% vs 6.45%, *p*=0.003) in the cilostazol-treated group was extremely significantly lower than that in the cilostazol-free group (Tab. 1-2 and Fig. 3). These results indicate that cilostazol treatment has a good preventive efficacy on the occurrence of PSD. It’s suggested that clinicians can use cilostazol to prevent further exacerbation of depression if they observe that AIS patients (no cilostazol contraindications history) have a depressive tendency during medical practice.

**Table 2.**
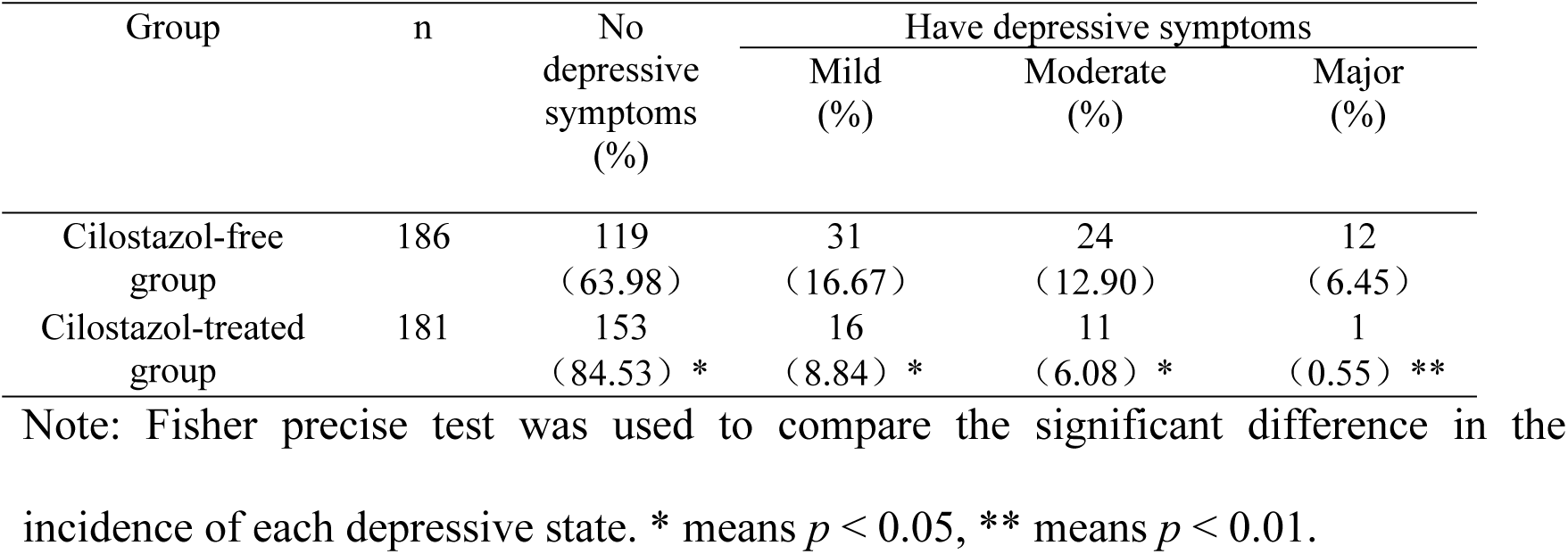
The incidence of PSD in the two groups at the end of follow-up.

| Group | n | No depressive symptoms (%) | Have depressive symptoms |  |  |
| --- | --- | --- | --- | --- | --- |
|  |  |  | Mild (%) | Moderate (%) | Major (%) |
| Cilostazol-free group | 186 | 119<br>(63.98) | 31<br>(16.67) | 24<br>(12.90) | 12<br>(6.45) |
| Cilostazol-treated group | 181 | 153<br>(84.53) * | 16<br>(8.84) * | 11<br>(6.08) * | 1<br>(0.55) ** |

Note: Fisher precise test was used to compare the significant difference in the incidence of each depressive state. \* means $p < 0.05$ , \*\* means $p < 0.01$ .

**Figure 3.**
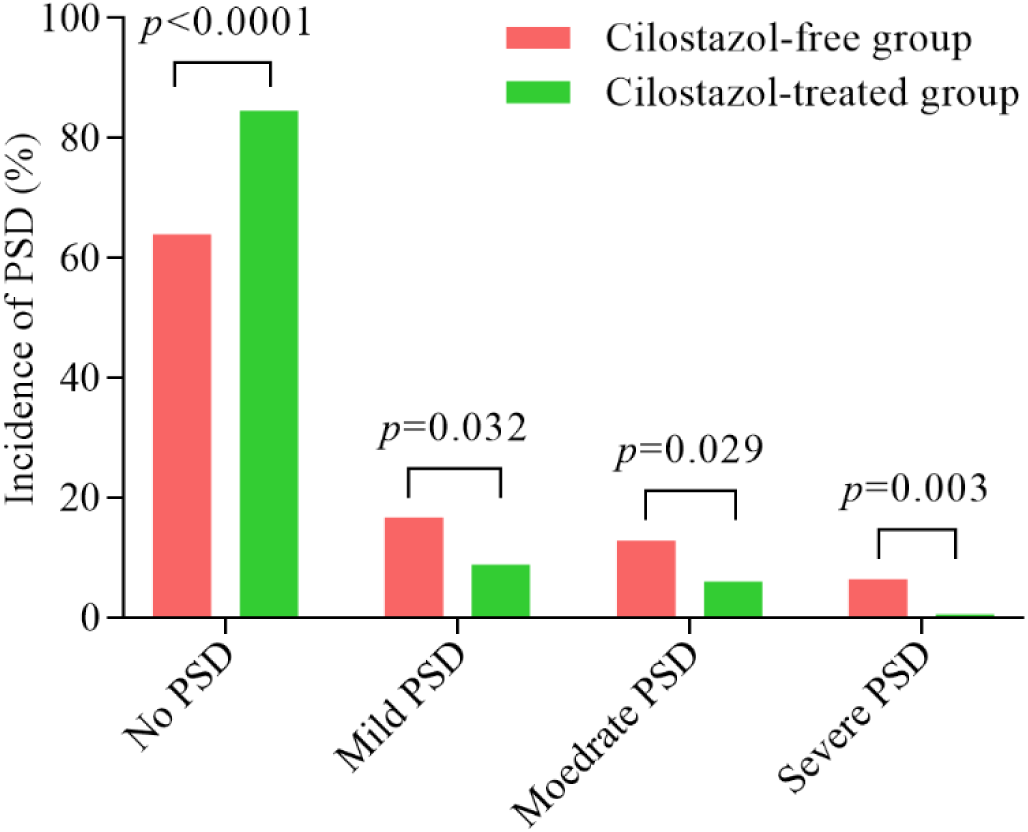
The incidence of PSD in patients of the cilostazol-free group and cilostazol-treated group at the end of follow-up. Fisher precise test was used to compare the significant difference in the incidence of each depressive state.

We also analyzed the correlation between the NIHSS scores and the HAMDS scores in the two groups at the end of follow-up, and the results showed that there was a positive correlation between the HAMDS scores and the NIHSS scores in the two groups, but the correlation between the HAMDS scores and the NIHSS scores in the cilostazol-treated group(R^2^=0.144) was lower than that in the cilostazol-free group (R^2^=0.308) (Fig. 4). These results suggest that cilostazol is more effective in preventing

**Figure 4.**
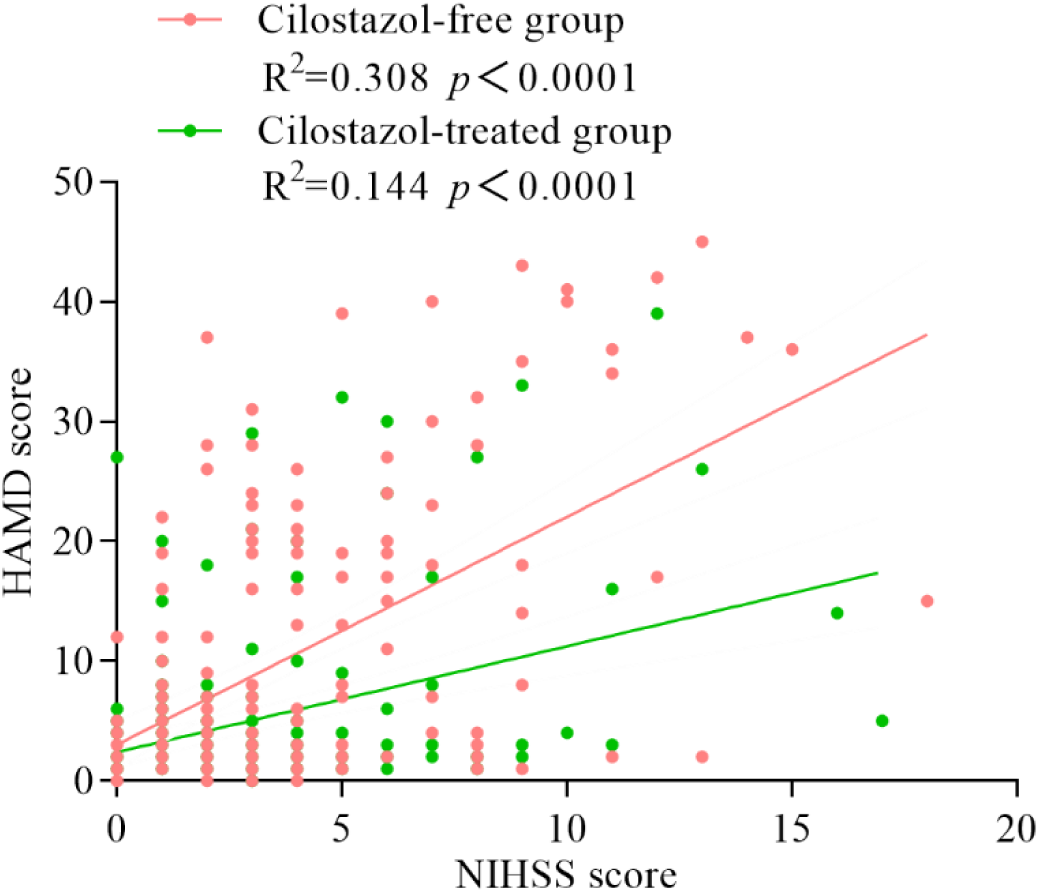
Correlation analysis between NIHSS score and HAMDS score in patients of the cilostazol-free group and cilostazol-treated group at the end of follow-up. Spearman rank correlation test was used to analyze the correlation between NIHSS score and HAMDS score. R^2^ is the determining coefficient. The higher R^2^ is, the stronger the correlation between NIHSS score and HAMDS score is. *p* < 0.0001 indicates that the correlation coefficient R has extremely significant statistical significance.

PSD than in improving AIS itself. These results also suggest that when facing AIS patients with depressive tendency, adjuvant therapy with cilostazol may have a better clinical effect than other conventional drugs, and the rehabilitation degree of nerve defect will be significantly improved, while the incidence of PSD will be significantly reduced.

### Hs-CRP and HCY

Others and our previous study showed that Hs-CRP and HCY can be used as predictive markers for the occurrence of PSD[29, 30]. Therefore, we examined the plasma levels of Hs-CRP and HCY in the two groups of patients at admission and at the end of follow-up, respectively, to verify the therapeutic efficacy of cilostazol on PSD and AIS from the physiological perspective. The results showed that the levels of Hs-CRP and HCY in the two groups were close at admission, while at the end of follow-up, the levels of Hs-CRP (*p*=0.046) and HCY (*p*=0.031) in patients of cilostazol-treated group were significantly lower than those in the cilostazol-free group (Tab. 1 and Fig. 5). The results also support that cilostazol treatment has a good preventive efficacy on the occurrence of PSD.

**Figure 5.**
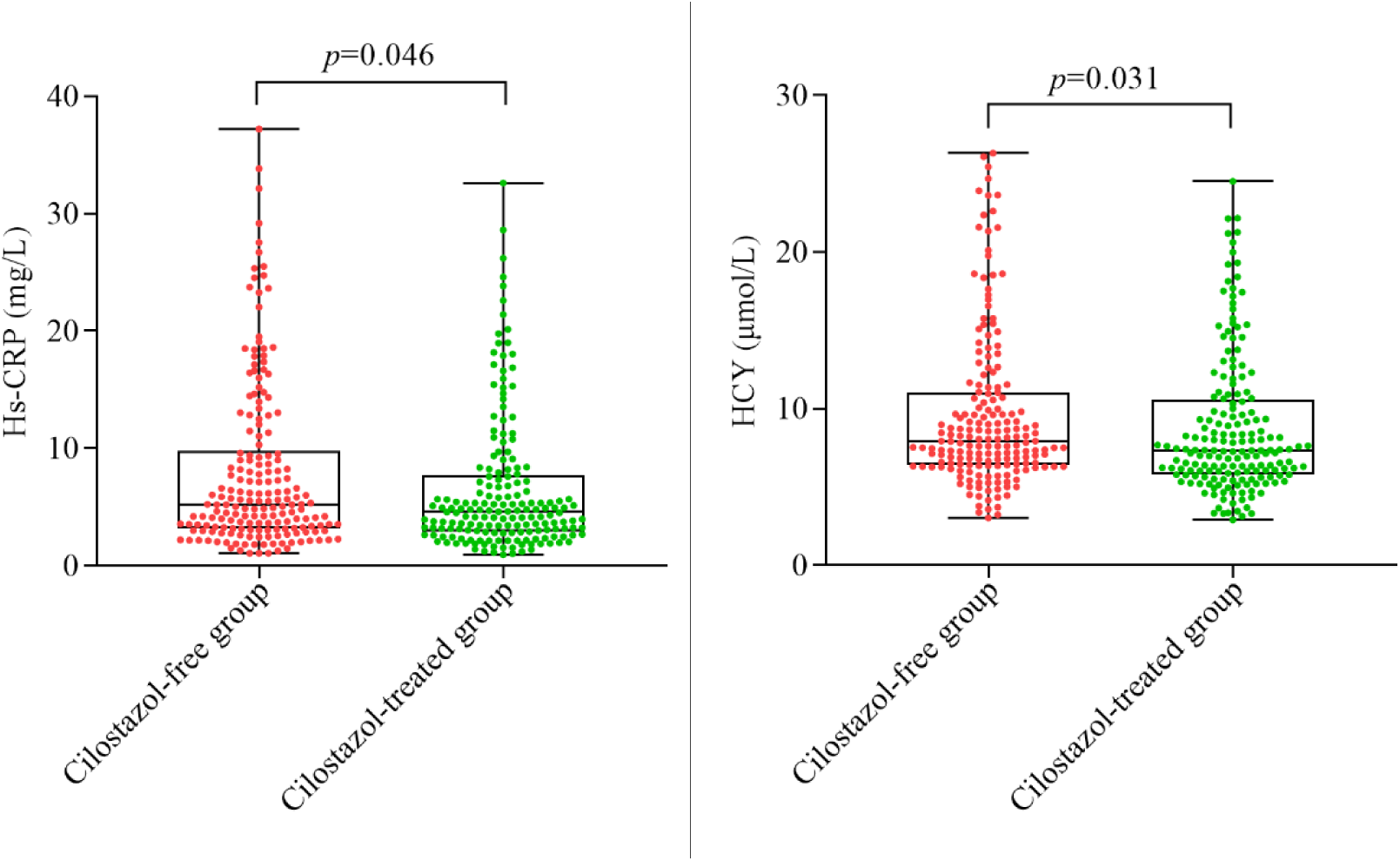
Plasma Hs-CRP and HCY levels in patients of the cilostazol-free group and cilostazol-treated group at the end of follow-up. Mann-Whitney *U* test was used to analyze whether there was a significant difference in Hs-CRP and HCY levels between the two groups. The upper and lower edge of the box represents the upper quartile and the lower quartile.

### TNF-α, IL-1β and NLR

Both depression and PSD are related to the level of inflammation[1, 11–13, 31], and cilostazol has been proven to improve the level of inflammation[8–10, 32]. Therefore, we further examined the plasma levels of TNF-α, IL-1β and NLR in the two groups of patients at admission and at the end of follow-up, respectively, to verify whether the preventive effect of cilostazol on PSD is related to its inflammatory regulation. The results showed that there were no significant differences in the levels of TNF-α (*p*=0.341), IL-1β (*p*=0.199) and NLR (*p*=0.375) between the two groups at admission (Tab. 1), while at the end of follow-up, the levels of TNF-α (*p*=0.025), IL-1β (*p*=0.028), and NLR (*p*=0.017) were significantly lower in patients of cilostazol-treated group than in those of cilostazol-free group (Fig. 6). These results indicate that cilostazol may play an important role in PSD prevention through its anti-inflammatory efficacy. The mechanism of cilostazol preventing PSD may be related to its anti-inflammatory effect. Clinicians can also predict the possibility of AIS patients developing to PSD by observing the duration and levels of Hs-CRP, HCY, TNF-α, IL-1β or NLR indicators in AIS patients, so as to take appropriate countermeasures.

**Figure 6.**
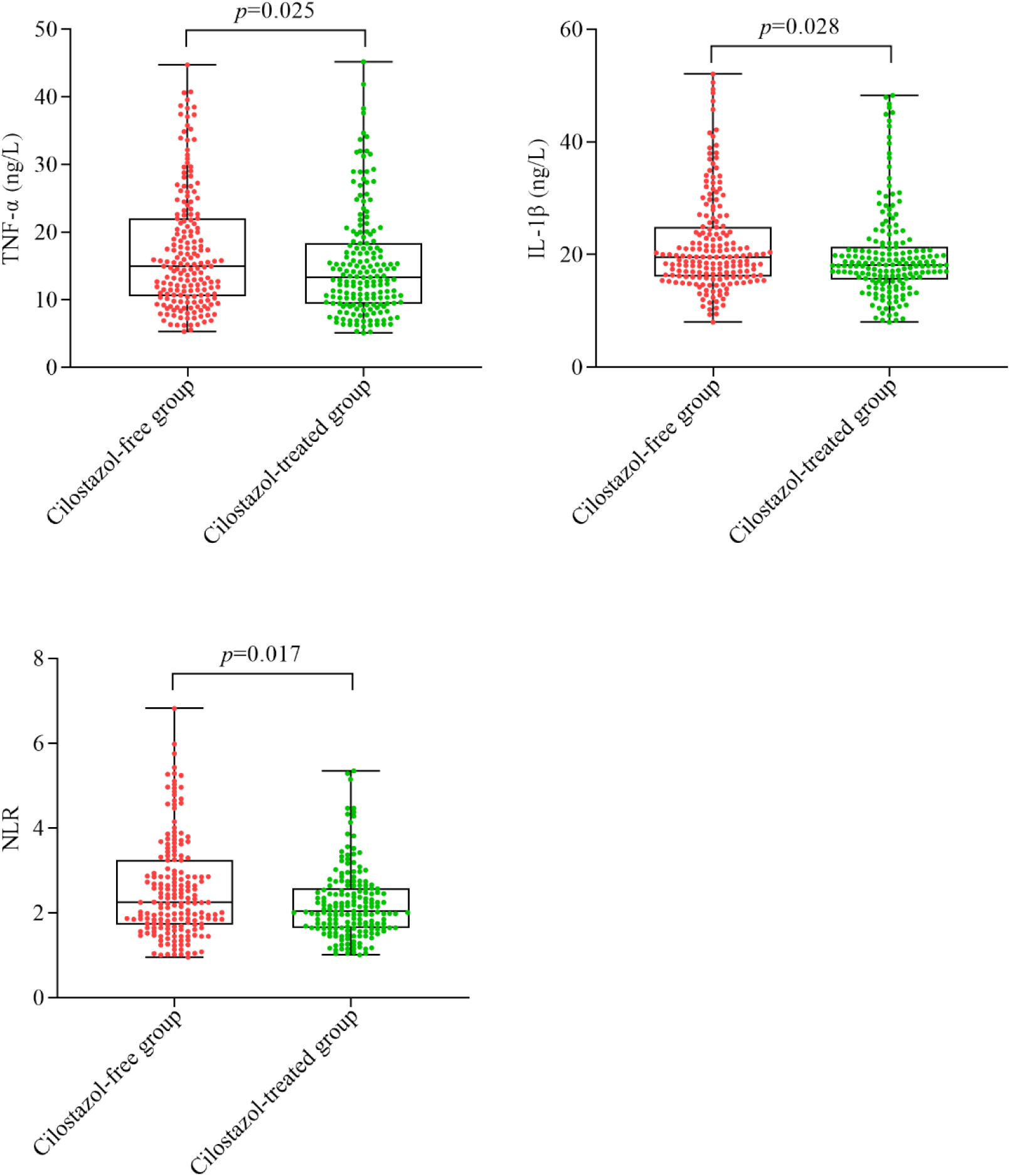
Plasma TNF-α, IL-1β and NLR levels in patients of the cilostazol-free group and cilostazol-treated group at the end of follow-up. Mann-Whitney *U* test was used to analyze whether there were significant differences in the three indexes between the two groups. The upper and lower edge of the box represents the upper quartile and the lower quartile.

## Discussion

The therapeutic efficacy of cilostazol on IS and the prevention of secondary recurrence have been confirmed, therefore, it has been used as a therapeutic drug for IS in Asian population. In the clinical treatment of IS, there have been case reports that cilostazol may prevent the occurrence of PSD, however, there is no systematic and rigorous clinical trial and follow-up evidence[18–22]. In order to fully confirm the preventive efficacy of cilostazol on PSD in Asian population, 431 AIS patients were recruited at Xinxiang First People’s Hospital from 2021 to 2022, follow-up and diagnostic analysis were conducted for 6 months, NIHSS and 24-items HAMDS scores, the plasma levels of Hs-CRP, HCY, TNF-α, IL-1β and NLR were measured.

NIHSS score showed that at the end of follow-up, patients in the cilostazol-treated group was significantly lower than those in the cilostazol-free group, confirmed that cilostazol has a good therapeutic efficacy on IS. HAMDS scores showed that at the end of follow-up, the incidence of PSD in cilostazol-treated group was much lower than that in cilostazol-free group, suggesting that cilostazol had a preventive efficacy on the occurrence of PSD. We also compared the correlation between NIHSS scores and HAMDS scores between the two groups, and found that cilostazol significantly reduced the correlation between NIHSS scores and HAMDS scores, which indicated cilostazol is more effective in preventing PSD than in improving AIS.

It has been reported that the occurrence of PSD is often related to the infarcted brain area of stroke patients, and patients with infarcted brain areas in frontal cortex, parietal lobe, occipital lobe, temporal lobe and hippocampus are more likely to show cognitive impairment in the process of stroke prognosis[33–36]. At admission, we found that there was no significant difference in infarcted brain area between the two groups, so the incidence of PSD was similar between them (Tab.1). However, at the end of follow-up, the incidence of PSD in patients of cilostazol-treated group was significantly lower than that in patients of cilostazol-free group (Tab.2), which supported the preventive efficacy of cilostazol on PSD. Clinical studies have also found that cilostazol has the effect of improving the cognitive status and reducing the recurrence rate in AD and IS patients[5, 20, 37–39], which is consistent with our results. In our study, the incidence of adverse drug event was also measured between patients of cilostazol-treated group and those of cilostazol-free group, and there was no difference between the two groups, suggesting that cilostazol is feasible for the prevention of PSD.

Others and our previous study found that plasma Hs-CRP and HCY levels could be used as markers of PSD[29, 30]. Therefore, in this study, we further detected the plasma Hs-CRP and HCY levels of all patients to verify the preventive effect of cilostazol on PSD from the perspective of biomarker molecules. The results showed that the mean levels of Hs-CRP and HCY in patients of cilostazol-treated group were significantly lower than those in patients of cilostazol-free group, which was consistent with the results of NIHSS and HAMDS scores. When discussing the therapeutic efficacy of cilostazol on IS patients, Saji et al. found that cilostazol can effectively reduce the plasma levels of Hs-CRP, MMPs and IL-6 in IS patients, which is conducive to reducing inflammatory related progressive stroke brain injury[40]. Saji’s results support our findings.

With the development of the inflammatory hypothesis of depression, the degree of deviation from normal plasma levels of TNF-α, IL-1β and NLR in patients has been proved to be positively correlated with the severity of PSD[11–13, 41–43]. TNF-α in peripheral blood of PSD patients not only participates in peripheral inflammatory response, but also mediates the injury of nerve cells, and aggravates the cognitive impairment of PSD patients by interfering with the plasticity of neuronal synapses[11, 43]. In our study, the mean level of TNF-α in peripheral blood of patients of cilostazol-treated group was significantly lower than that of patients of cilostazol-free group, which suggests that the reduced incidence of PSD and favorable prognosis of stroke in patients of cilostazol-treated group may be related to the decreased expression of TNF-α.

IL-1 was first identified as a neuropeptide cytokine, which can be produced by endothelial cells, microglia, astrocytes and neurons in the peripheral and central nervous system. Under normal circumstances, only a small amount of IL-1 is expressed in the brain, which is mainly distributed in the hippocampus, hypothalamus and cortex. After cerebral ischemia-reperfusion injury, IL-1 is significantly increased, and the increase amplitude is related to the degree and duration of ischemia[12, 13]. IL-1β is a member of the IL-1 family, when IS occurs, the level of IL-1β will increase significantly within 24 hours[44]. Owen found that plasma levels of IL-1β were significantly increased in patients with major depression compared with healthy people[45]. Murata et al. reported that elevated levels of IL-1β were closely related to refractory bipolar depression[12]. A survey of 305 IS patients showed that homeostatic model assessment of insulin resistance (HOMA-IR) and IL-1β were closely related to the occurrence of PSD, and these two factors together improve the ability to assess early PSD[13]. On the basis of these studies on the association between IL-1β and PSD, we detected the expression changes of plasma IL-1β in the two groups of patients at admission and at the end of follow-up, and found that the expression changes of IL-1β were similar to TNF-α. Combined with the results of HAMDS score and stroke prognosis, we concluded that cilostazol could prevent the occurrence of PSD by reducing the level of inflammation in peripheral and central nervous system.

Clinical studies in the past two years have shown that NLR is independently associated with the occurrence of PSD and can be used as a predictor of PSD[41, 42, 46]. Hu et al. retrospectively analyzed the clinical data of 376 patients with first episode IS in the First Affiliated Hospital of Wannan Medical College (Yijishan Hospital) and pointed out that NLR≥4.02 was independently associated with the occurrence of PSD at 6 months after stroke, which has guiding significance in clinical screening of early PSD[42]. Chen et al.’s clinical trial (180 healthy controls and 299 IS patients) showed that NLR≥3.70 was independently associated with the occurrence of PSD one month after stroke[46]. According to the current research results[41, 42, 46, 47], we surmise the explanation for the elevated NLR in PSD patients is as follows: After stroke, the BBB of patients is damaged to different degrees[47], the permeability of BBB changes, and neutrophils and lymphocytes in the peripheral circulation enter the central nervous system through the damaged BBB, inducing intracranial inflammation. The increased level of intracranial inflammation will release neurotransmitters in a variety of emotional regulatory networks in the brain, and this effect will accumulate and eventually induce the occurrence of mental disorders or PSD[41, 42, 46]. In view of its important reference value, we calculated the NLR of the two groups of patients at admission and at the end of follow-up, and found that the average NLR in patients of cilostazol-treated group was significantly lower than that of patients of cilostazol-free group, which suggests that cilostazol effectively improves inflammatory cells activity of IS patients, reduces its harassment of the nervous system, and prevents the occurrence of PSD.

After two years of persistent case collection, based on the analysis of Hs-CRP and HCY, which can simultaneously characterize stroke and depression, and the analysis of TNF-α, IL-1β and NLR, which can simultaneously characterize the peripheral and central systems, as well as the NIHSS and HAMDS scores of AIS patients, we systematically investigated the efficacy of cilostazol in the treatment of stroke, depression and inflammation, and found the levels of these biomarker molecules, NLR, and NIHSS and HAMDS scores in patients of cilostazol-treated group were all significantly lower than those of cilostazol-free group. Our results completely demonstrate the good therapeutic effect of cilostazol on AIS and the excellent preventive effect on PSD. From the perspective of PSD prevention efficacy, cilostazol is a better therapeutic agent for AIS.

Studies have shown that both stroke and depression patients are accompanied by elevated levels of inflammation to some extent[1, 11–16]. Our findings suggest that the mechanism by which cilostazol reduces the incidence of PSD in AIS patients may be related to its regulation of the inflammatory state in AIS patients. In addition to the anti-platelet aggregation and vasodilator effects of clinical application, cilostazol has been proved to have anti-inflammatory effects in major depressive disorder related clinical experiments and model rats with hypercholesterolemia, diabetes, and colitis. It has good inhibitory effect on NF-κB, TNF-α, IL-6 and IL-1β[8, 18, 48, 49]. We speculate that cilostazol can effectively inhibit thrombosis and block blood vessels in patients with AIS through anti-platelet aggregation and vasodilation, and then further inhibit the inflammatory response in the focal area caused by stroke, inhibit the progression of AIS, and effectively prevent the recurrence of AIS. The persistent inflammatory state after AIS is closely related to the occurrence of PSD[1, 13–16]. We speculate that cilostazol effectively prevents the occurrence of PSD by continuously disrupting the progress of AIS to PSD. This may also be the reason why we observed a lower correlation between NIHSS scores and HAMDS scores in the cilostazol-treated group (Fig. 7). The specific mechanism of cilostazol in regulating the level of inflammation and preventing PSD will be further discussed in subsequent animal experiments. These results also suggest that if clinicians observe that the inflammation-related indexes such as TNF-α, IL-1 β or NLR are abnormally high, or the prognosis of the patient is not good, and the NIHSS score does not achieve the expected therapeutic effect, they should pay more attention to the emotional changes of the patient, observe whether the patient has depression-related symptoms, and whether antidepressant drugs need to be added to assist the treatment of stroke.

**Figure 7:**
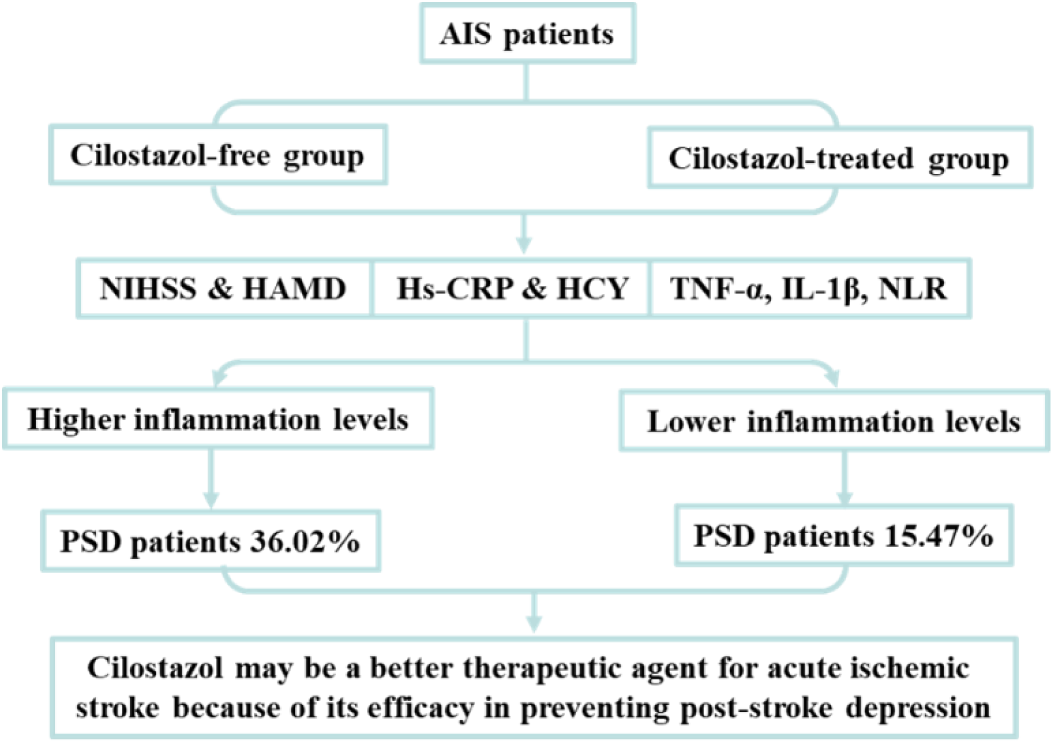
The potential mechanisms of cilostazol on PSD prevention.

In addition, the similarities and differences between cilostazol and the other anti-inflammatory drugs in the anti-inflammatory mechanism in clinical PSD precaution have not been reported in detail, whether ordinary anti-inflammatory drugs can also reduce the incidence of PSD and improve the prognosis of stroke patients needs further experimental investigation. In addition, although fluoxetine, a commonly used antidepressant, reduces the incidence of PSD, it is not recommended for the prevention of PSD. Because fluoxetine increases the risk of falls, bone fractures and epileptic seizures and hyponatremia in stroke patients[3, 4]. We expect more researchers to pay attention to the study of cilostazol and other anti-inflammatory drugs on PSD prevention, and work together to find a feasible treatment plan for PSD prevention.

## Limitations

This study also has the following limitations: Firstly, PSD is a disease of the central nervous system, which is separated from the periphery by the blood-brain barrier. Therefore, brain biopsy should be the most convincing way for our indicators. While, brain biopsy is obviously not feasible. Secondly, some of the cases we collect are from rural areas and others are from cities. These families have different economic conditions and degrees of burden on the disease, which will have different effects on the mood of patients. However, we did not analyze the economic situation of patients, and this confounding factor may have a certain bias on the results. Thirdly, the rehabilitation of AIS patients and the occurrence of PSD are closely related to the care they receive. Although our clinicians have advised the family members of each patient, we did not provide unified training in nursing-related knowledge to all patients’ families so that this universal confounding factor can be effectively controlled. In future research, we will use further rigorous experimental design to reduce the impact of these potential confounding factors on the validity and generalizability of experimental findings.

In addition, because stroke is a complex disease with high recurrence rate and relatively poor prognosis, the probability of combining drugs is extremely high. This situation bring great challenges to the further profound verification of the potential efficacy of cilostazol in preventing PSD and its popularization and application in clinic. We expect more powerful researchers to discuss this topic more deeply.

## Data Availability

7785

## Authors’ contributions

CZ Tang conceived of the study. YL Zhang, GG Yang, R Wang, X Wang, XY Tan and CZ Tang initiated the study design and YL Zhang, R Wang, X Wang, XY Tan helped with implementation. CZ Tang and R Wang are grant holders. YL Zhang and GG Yang provided statistical expertise in clinical trial design and YL Zhang is conducting the primary statistical analysis. All authors contributed to refinement of the study protocol and approved the final manuscript.

## ACKNOWLEDGEMENTS

We thank all of the patients who participated to this study, the clinical teams from the department of Neurology at Xinxiang First People’s Hospital. And also, we thank the reviewers for helpful comments on the manuscript. In addition, we thank the grant from Scientific and Technological Project of Henan Province (No.212102310836, No.232102310110) and Scientific and Technological Project of Xinxiang City (No. GG2019002).

